# Epilepsy Lateralization by Elastic Net Regularization of a Diffusion Tensor Imaging Biomarker using Machine Learning

**DOI:** 10.64898/2026.11.03.26367189

**Authors:** Rebecca J. Lepping, McKenzie Pearl, Xi Chen, Jianxing Hu, Yanming Li, Murshalina Akhter, Mark Farrenburg, Patrick Landazuri, Caleb M. Pearson

## Abstract

**Objective:** Accurate lateralization of the epileptogenic focus is essential for presurgical evaluation of patients with drug-resistant epilepsy. This study examined whether diffusion tensor imaging (DTI) measures of white matter microstructure, combined with clinical and demographic variables, could classify epilepsy lateralization using machine learning.

**Methods:** We present a retrospective analysis of 334 adult patients undergoing presurgical epilepsy evaluation at the University of Kansas Health System. Fractional anisotropy (FA) and mean diffusivity (MD) were extracted from 36 bilateral white matter tracts and combined with clinical and demographic variables. Several supervised machine learning approaches were evaluated, with elastic net logistic regression selected for detailed analysis. Models classified right-sided, left-sided, and bilateral or generalized epilepsy. Performance was evaluated with area under the receiver operating characteristic curve (AUC), accuracy, sensitivity, and specificity as performance metrics.

**Results:** Elastic net models demonstrated good discrimination across classification tasks, with AUC values ranging from 0.76 to 0.88. The strongest performance distinguished left-sided epilepsy from bilateral or generalized epilepsy (AUC = 0.88; accuracy = 0.90; sensitivity = 0.90; specificity = 0.89). Discrimination between unilateral epilepsy and bilateral or generalized epilepsy was also good (AUC = 0.83). Classification of right-sided versus left-sided epilepsy demonstrated lower performance (AUC = 0.76).

**Significance:** DTI-derived white matter features analyzed using a machine learning approach may provide complementary information for epilepsy lateralization during presurgical evaluation. Further studies incorporating other pre-surgical evaluation data into the machine learning model are needed to establish the clinical utility of this approach.

**Key Points:**

- A machine learning model was tested using diffusion tensor imaging and clinical data to retrospectively classify epilepsy lateralization.
- Our model balanced sensitivity and specificity in epilepsy localization, suggesting potential as an interpretable surgical decision tool.
- Findings support the hypothesis that white matter tracts contain useful information about seizure propagation to distal brain regions.
- Further model validation is needed before this can be used as a clinical tool.

---

Epilepsy is a prevalent neurological disorder, affecting around 50 million people worldwide or approximately one percent of the world’s population [1, 2]. According to the International League Against Epilepsy, seizures are defined as “transient occurrence of signs and/or symptoms due to abnormal excessive synchronous neuronal activity of the brain” [3]. Epilepsy is characterized by recurrent, unprovoked seizures that manifest in a variety of ways depending on where the seizure has originated in the brain and its network [2].

Epilepsy can impose potential lifelong impacts on physical and mental health while significantly limiting quality of life due to associated social stigma, potential medical expenses, and functional limitations on daily living and lifestyle [2]. Antiseizure medications (ASM) are the first line treatment for epilepsy; however, approximately one third of patients continue to have seizures despite adequate ASM treatment and have drug resistant epilepsy (DRE) [4]. Those with DRE may benefit from surgical treatment.

The presurgical evaluation for epilepsy is well documented and begins with video electroencephalogram (vEEG) to help localize the seizure focus. This data is used in conjunction with imaging studies such as 3 Tesla (3T) magnetic resonance imaging (MRI), magnetoencephalography (MEG), positron emission tomography (PET), ictal single photon emission computed tomography (SPECT) or high-field (7T) MRI to plan for surgery. For some patients this provides sufficient information to proceed with a therapeutic surgery. Still, others are not localized or lateralized sufficiently necessitating more invasive studies like stereo electroencephalogram (sEEG).

Despite the high diagnostic yield of conventional MRI, a common reason for proceeding to sEEG is that a large minority of individuals with epilepsy have no apparent epileptogenic lesion. For instance, approximately 20-30% of temporal lobe epilepsy patients present with no visible abnormalities on epilepsy protocol MRI scans [5–7]. Advanced MRI techniques such as functional MRI (fMRI) and diffusion tensor imaging (DTI) can complement structural MRI data by mapping critical functional areas of the brain, providing additional insight into potential network dysfunction, aid in lateralization and localization of the epileptogenic zone (EZ) and assist in presurgical planning. Presurgical brain mapping seeks to improve surgical targeting and outcomes by identifying white matter structural changes that potentially signify the presence of epilepsy and subsequently and ideally help either lateralize the EZ to the right or left hemisphere or identify a generalized or bilateral source [8, 9]. Further, being able to identify seizure propagation networks with tractography could allow targeting of involved networks with neuromodulation devices such as responsive neurostimulation and deep brain stimulation.

DTI is a MRI based imaging technique that maps water molecule diffusion in multiple directions along white matter tracts enabling visualization of underlying tissue architecture and connectivity [10]. White matter tracts consist of axon bundles that connect different cortical regions to facilitate rapid neuronal communication. DTI models like DTI tractography (DTI-T) can be used to identify subtle white matter abnormalities within the epileptic network to help plan surgical trajectories that minimize disruption to critical pathways [10]. Two principal DTI-derived metrics consistently linked to microstructural abnormalities in epilepsy are Fractional Anisotropy (FA) and Mean Diffusivity (MD). FA ranges from 0=spherical to 1=linear and captures the degree of directionality of water diffusion and serves as a marker of the white matter tract coherence and integrity [5]. The MD is the overall magnitude of water diffusion in all directions. MD is highest in free water (MD=1) and provides an index of cellular density and tissue integrity. A reduced FA and increased MD are common hallmarks of the EZ, even when MRI is “normal” [5].

Recent advances in machine learning have significantly impacted the field of epilepsy by allowing researchers and clinicians to leverage algorithms, large datasets, and advanced data analysis techniques to aid in prediction, classification and detection of seizures [1]. Our aim was to determine if epilepsy source lateralization could be determined from a combination of pre-surgical DTI and clinical variables using a statistical machine learning model. We conducted a retrospective phase 2: preliminary biomarker validation study [11] to determine whether DTI-T metrics of FA and MD extracted from individual white matter tracts improve classification of patients’ seizure lateralization from consensus clinical conference diagnosis. We hypothesize that machine learning models that include DTI-T biomarker metrics will yield accurate, sensitive, and specific clinical classification of patients’ EZ as originating in the left hemisphere, right hemisphere, or having a general or bilateral source.

## Methods

### Participants and Recruitment

In this retrospective phase 2: preliminary biomarker validation study [11], adult epilepsy surgical candidates (>18 years of age) were referred for functional brain mapping fMRI and DTI as part of their epilepsy pre-surgical workup with the University of Kansas Health System.

Candidates were excluded from DTI mapping if a brain mass was identified during clinical review, if they had a vagus nerve stimulator (VNS), or if the scan could not be completed for any reason.

### Standard Protocol Approvals, Registrations and Patient Consent

This study was approved by the University of Kansas Medical Center Human Research Protection Program, and all data were collected into the research database from retrospective chart review as part of an ongoing approved IRB Study (Approval # STUDY00160649) with waiver of consent in accordance with the Declaration of Helsinki. All patients at the University of Kansas Medical Center sign a general consent for retrospective chart review. Demographic and clinical variables extracted from the medical records included gender, race, ethnicity, handedness, age at imaging, and education level. Pre-surgical DTI-T imaging biomarker data, seizure lateralization from consensus clinical conference diagnosis, surgery status or type, and availability of pre- and post-neuropsychological testing were also obtained from the medical records. No other post-surgical outcome data are included in our study. Our manuscript follows the GREENBEAN Checklist for reporting evaluations of neurophysiological data [11]. The checklist is included in supplemental information (Table S1).

### MRI and DTI Data Acquisition

MRI data were acquired using a Siemens (Erlangen, Germany) Skyra 3T MRI scanner with a 16-channel head coil. The high-resolution structural brain images were acquired using a three-dimensional T1-weighted magnetization prepared rapid acquisition gradient echo (MPRAGE) sequence with TE/TR/inversion time of 2.54/1900/900 ms and voxel size of 0.5×0.5×1.00 mm. Diffusion tensor images (DTI) were collected with 28 axial slices, TE=95 ms, and TR=3800 ms. A DTI diffusion scheme was used, and a total of 30 diffusion sampling directions were acquired with b-values of 0 and 1000 s/mm2 with three averages. The in-plane resolution was 1.875 mm, FOV = 240 mm, base resolution = 128. The slice thickness was 4.0 mm. GRAPPA acceleration factor = 2 was applied.

### DTI Processing and Analysis

DTI data were processed and analyzed using DSI Studio [12], the Hou version [13] using software-recommended settings for automated fiber tracking. Briefly, the restricted diffusion was quantified using restricted diffusion imaging [14]. The diffusion data were reconstructed using generalized q-sampling imaging [15] with a diffusion sampling length ratio of 1.25. A deterministic fiber tracking algorithm [16] was used. A seeding region was placed on the selected tract. The anisotropy threshold was 0.204657. The angular threshold was randomly selected from 15 degrees to 90 degrees. The step size was randomly selected from 0.5 voxel to 1.5 voxels. Tracks with length shorter than 30 or longer than 300 mm were discarded. A total of 50000 seeds were placed.

Thirty-six anatomic white matter tracts were identified for analysis for the purposes of the study (see supplemental information Tables S2 and S3) based on their significance to the brain structure and function. DTI-T predictors were derived from bilateral white matter tracts and included diffusion metrics from major association, projection, and limbic pathways. For our study, emphasis was placed on tract-level FA and MD measurements from homologous left and right tracts, capturing potential microstructural asymmetry associated with epileptogenic networks. Representative pathways included the arcuate fasciculus, cingulum, frontal and parietal aslant tracts, inferior fronto-occipital fasciculus, inferior longitudinal fasciculus, middle longitudinal fasciculus, superior longitudinal fasciculus subdivisions, uncinate fasciculus, corticostriatal tracts, thalamic radiations, fornix, vertical occipital fasciculus, and cerebellar pathways.

### Statistical Modeling

The outcome for this study was defined as lateralization of the EZ to the right hemisphere, left hemisphere, or bilateral hemispheres indicating a generalized localization or bilateral epilepsy, determined from the consensus clinical conference diagnosis that did not consider information from DTI assessment. Pairwise binary classifiers were constructed between predefined lateralization classes (right vs left, right vs bilateral, left vs bilateral, and right + left vs bilateral), allowing discrimination among clinically meaningful lateralization categories. Predictor variables comprised demographic and clinical characteristics together with quantitative DTI-T tract-based diffusion biomarker features as described above.

All analyses were conducted in R (v. R-4.5.1) [17]. Raw variables were inspected prior to modelling. Categorical predictors were converted to factor variables. Missing values in categorical fields were treated as an explicit “Missing” category, whereas missing numeric values were imputed using the median value estimated from the training data only. Continuous predictors were standardized by centering and scaling. Categorical variables were transformed using one-hot encoding to generate model-ready design matrices [18]. Analysts were not blinded to clinical information.

### Model Development

A variety of machine learning models exist. Logistic regression is a generalized linear modelling approach used when the outcome variable is binary [19]. However, logistic regression may be unstable when the number of predictors is large or when substantial multicollinearity exists among covariates. This is particularly relevant in epilepsy data, so five supervised machine learning approaches were evaluated, including support vector machine (SVM), random forest (RF), gradient boosting machine (GBM), deep neural network (DNN), and elastic net-regularized logistic regression. These algorithms were selected to represent different modelling strategies, including linear regularized regression, kernel-based classification, ensemble learning, and deep learning, allowing comparison of their ability to capture linear and nonlinear relationships between DTI-derived features, clinical covariates, and epilepsy lateralization [20–23]. All models were evaluated across four binary classification tasks using stratified training (80%) and testing (20%) datasets. Model performance was assessed using accuracy, sensitivity, specificity, and area under the receiver operating characteristic curve (AUC). Among these candidates, the elastic net logistic regression was selected as the best model for further analysis because it achieved the highest mean AUC across the four tasks and demonstrated a more balanced sensitivity and specificity profile compared with the alternative approaches.

Elastic net is a penalized regression method that combines the strengths of lasso and ridge regression by simultaneously applying L1 and L2 regularization [24]. Elastic net is particularly advantageous in datasets with correlated features or when the number of predictors is large relative to the sample size. Because elastic net is a linear and intrinsically interpretable model, the most predictive variables can be quantified using the magnitude of standardized regression coefficients. Features with larger absolute coefficients are interpretable as stronger contributors to prediction, making elastic net an ideal model for this analysis aimed at not only classification accuracy, but also identifying clinically relevant predictors from within the model outcome. Elastic net logistic regression initially incorporated all available predictors without prior feature selection, allowing regularization to determine their contributions during model estimation. Features with larger absolute coefficients were considered more influential within the fitted multivariable model. However, these features may not be independently predictive, as overall classification performance depends on the combined information from all predictors that retained in the model.

### Training and Validation Strategy

For each classification task, the dataset was randomly partitioned into training (80%) and testing (20%) subsets using stratified sampling to preserve class proportions. Hyperparameter tuning was performed within the training set using repeated five-fold cross-validation (five repeats). This process systematically evaluates different combinations of model parameters to identify the configuration that optimizes predictive performance. Specifically, the elastic net mixing parameter (α), which balances L1 and L2 regularization, and the penalty parameter (λ), which controls the strength of regularization, were optimized by selecting the combination that achieved the highest mean cross-validated area under the receiver operating characteristic curve (AUC) [25].

To ensure reproducibility, fixed random seeds were specified for stratified training–test partitioning and cross-validation during hyperparameter tuning. These values were used consistently across the four binary classification tasks. For each binary classification task, patients were randomly allocated to training (80%) and held-out testing (20%) subsets using stratified sampling based on the outcome class to approximately preserve class proportions. To avoid information leakage between the training and held-out test sets, missing-value imputation and feature standardization parameters were estimated exclusively from the training data and subsequently applied to the corresponding test data.

### Prediction Threshold Selection

The elastic net model generated continuous predicted probabilities for each class. To derive binary class assignments, an operating threshold was selected using the Youden index, which maximizes the sum of sensitivity and specificity minus one [26]. This approach balances true positive and true negative classification for reporting diagnostic performance.

The equation of Youden index can be expressed as:

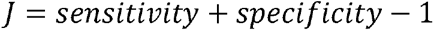

### Model Evaluation

Final model performance was assessed on the independent held-out test set. Metrics included accuracy, sensitivity, specificity, and area (AUC) under the receiver operating characteristic (ROC) curve. Receiver operating characteristic curves were generated for each pairwise classification task, and confusion matrices were used to summarize classification errors. Ranked coefficients were examined to identify the most informative diffusion and clinical variables associated with seizure focus classification. No corrections were made for multiple comparisons.

## Results

### Participant Characteristics

All 334 patients who completed pre-surgical DTI imaging data collection were included in the analysis. Participant demographics are described in Table 1. To compare the utility of DTI-T biomarkers for lateralization determination, our population included 131 patients (39%) with right hemisphere focal epilepsy, 157 (47%) with left hemisphere focal epilepsy, and 46 (14%) with bilateral or generalized epilepsy based on consensus clinical conference diagnosis (Table 2).

**Table 1.** Participant demographics.

| **Characteristic** | **N or Mean** | **Percent or SD** |
|---|---|---|
| Age | 38.52 | 13.28 |
| Gender |  |  |
| *Female* | 160 | 48% |
| *Male* | 174 | 52% |
| Race/Ethnicity |  |  |
| *African American* | 33 | 9.9% |
| *Asian/Pacific Islander* | 5 | 1.5% |
| *Caucasian* | 275 | 82.3% |
| *Hispanic* | 15 | 4.5% |
| *Hispanic/Caucasian* | 1 | 0.3% |
| *Native American* | 1 | 0.3% |
| *Other/Unknown* | 4 | 1.2% |
| Handedness |  |  |
| *Left* | 38 | 11.4% |
| *Right* | 295 | 88.3% |
| *Ambidextrous* | 1 | 0.3% |
| Education |  |  |
| *< 12 years* | 24 | 7.2% |
| *High School Diploma* | 99 | 29.6% |
| *Some College* | 108 | 32.3% |
| *Undergraduate Degree* | 47 | 14.1% |
| *Graduate Education* | 28 | 8.4% |
| *Unknown* | 28 | 8.4% |

Abbreviations: N – number of participants, SD – standard deviation.

**Table 2.** Clinical and surgical patient characteristics.

| **Characteristic** | **N** | **Percent** |
|---|---|---|
| Clinical Epileptic Zone Lateralization |  |  |
| *Right* | 131 | 39.0% |
| *Left* | 157 | 47.0% |
| *Bilateral or Generalized* | 46 | 14.0% |
| Surgery Type |  |  |
| *ATL* | 67 | 20.1% |
| *ATL+enceph* | 7 | 2.1% |
| *DBS* | 9 | 2.7% |
| *Frontal Resection* | 20 | 6.0% |
| *Insula Resection* | 2 | 0.6% |
| *LITT* | 9 | 2.7% |
| *Mesial Sparing ATL* | 6 | 1.8% |
| *Neocortical Temporal Resection* | 9 | 2.7% |
| *Occipital Resection* | 1 | 0.3% |
| *Parietal Resection* | 4 | 1.2% |
| *RFA* | 25 | 7.5% |
| *RNS* | 51 | 15.3% |
| *SLAH* | 22 | 6.6% |
| *VNS* | 3 | 0.9% |
| *No Surgery* | 99 | 29.6% |
| Pre-surgical Neuropsychological Testing |  |  |
| *Yes* | 303 | 91.0% |
| *No* | 27 | 8.1% |
| *Not available* | 4 | 1.2% |
| Post-surgical Neuropsychological Testing |  |  |
| *Yes* | 133 | 39.8% |
| *No* | 101 | 30.2% |
| *Not available* | 100 | 29.9% |

Abbreviations: N – number of participants, ATL – anterior temporal lobe, ATL+enceph – anterior temporal lobe and encephalocele, DBS – deep brain stimulation, LITT – laser interstitial thermal therapy, RFA – radiofrequency ablation, RNS – responsive neurostimulation, SLAH - stereotactic laser amygdalohippocampotomy, VNS – vagus nerve stimulation.

### Classification Prediction Modeling

The model performance of elastic net logistic regression model is shown in Table 3 and Figure 1. The elastic net model demonstrated good overall discrimination across all four of the paired binary epilepsy lateralization classification tasks, with AUC values of the independent 20% held-out test set ranging from 0.76 to 0.88 and accuracy across tests ranging from 0.71 to 0.90 (Table 3). Variance of the DTI-T biomarker across participants is shown in supplemental information (Figure S1).

**Figure 1.**
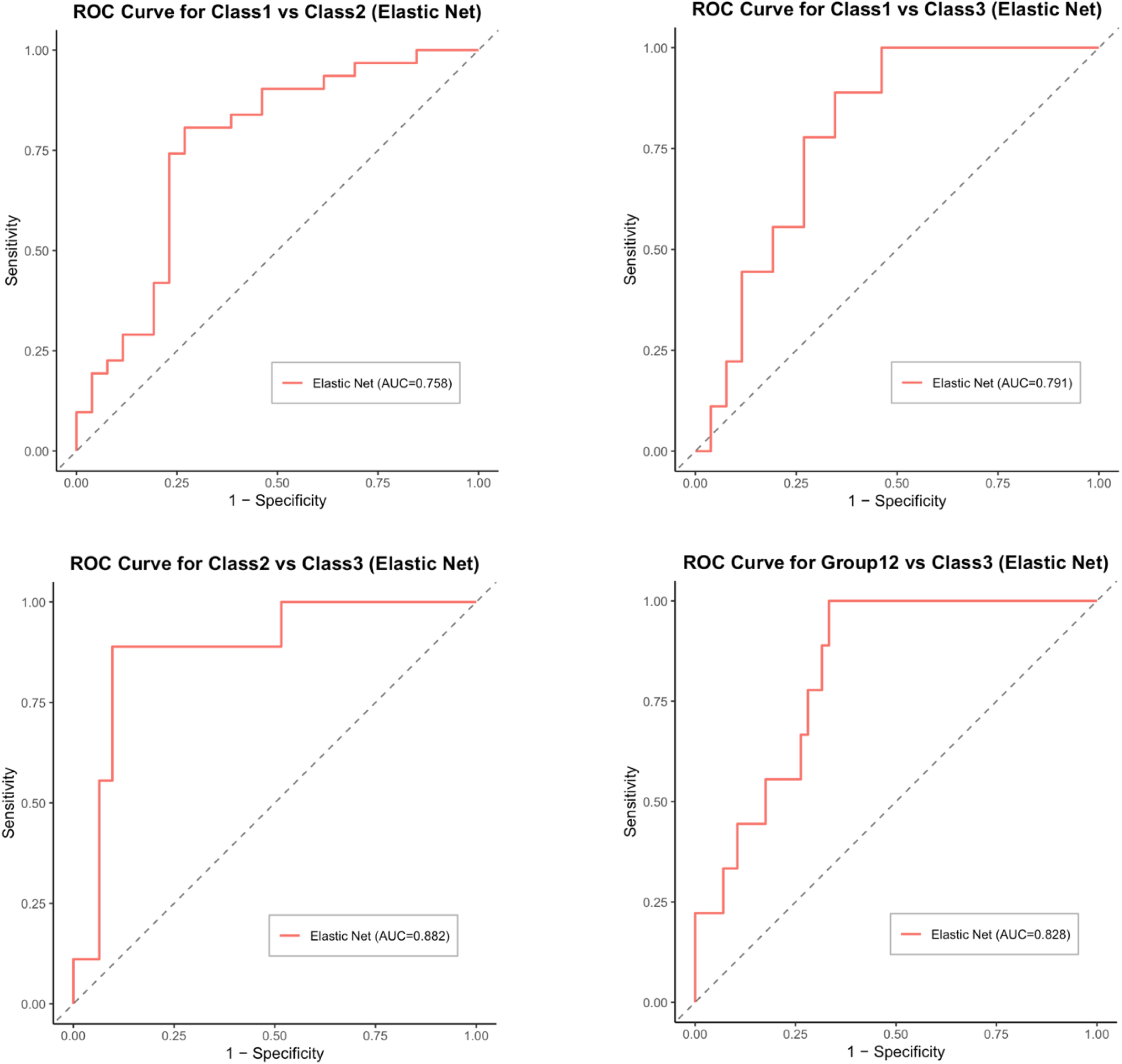
ROC curves comparing specificity and sensitivity for each of the four paired classification tasks on the held-out test set.

**Table 3.** Performance of the elastic net model on held-out test data.

| Tasks | Accuracy | Sensitivity | Specificity | AUC |
|---|---|---|---|---|
| Class 1 vs Class 2 | 0.77 | 0.73 | 0.81 | 0.76 |
| Class 1 vs Class 3 | 0.71 | 0.65 | 0.89 | 0.79 |
| Class 2 vs Class 3 | 0.90 | 0.90 | 0.89 | 0.88 |
| Class 1+2 vs Class 3 | 0.71 | 0.70 | 0.78 | 0.83 |

Note: Class 1 = right hemisphere; Class 2 = left hemisphere; Class 3 = bilateral or generalized; Class 1+2 = right + left hemisphere. Abbreviations: AUC – area under the curve.

The strongest performance was observed for classifying left hemisphere vs bilateral, which achieved an AUC of 0.88, accuracy of 0.90, sensitivity of 0.90, and specificity of 0.89, indicating that patients with left-sided focal epilepsy exhibited diffusion imaging patterns that were clearly distinguishable from those with bilateral or generalized epileptogenic involvement.

For left + right vs bilateral, the model also showed strong discrimination (AUC = 0.83) with balanced sensitivity (0.70) and specificity (0.78), suggesting that patients with unilateral focal epilepsy, regardless of side, could be reliably differentiated from those with bilateral or generalized seizure localization.

In contrast, lower right and left hemisphere discrimination was seen (AUC = 0.76), suggesting right and left sided focal epilepsy share partially overlapping microstructural patterns, as homologous left and right hemispheric tracts may undergo similar network remodeling despite opposite seizure laterality.

Moderate discrimination between right hemisphere and bilateral (AUC = 0.79) further supports the notion that bilateral/generalized epilepsy demonstrates broader diffusion abnormalities than right focal epilepsy, although overlap remained greater than for left-sided comparisons.

Figure 2 summarizes the common top predictive variables that were repeatedly selected by the elastic net models across the four pairwise classification tasks. This visualization provides some interpretabilities for the shared and task-specific predictors across tasks. Right sided vs left sided focal epilepsy had relatively weak differentiating effects, however, the strongest were the vertical occipital fasciculus and cingulum in the left hemisphere along with education level. Unilateral (right sided and left sided) versus bilateral classification had the strongest effects for various surgical outcomes in addition to DTI metrics and education.

**Figure 2.**
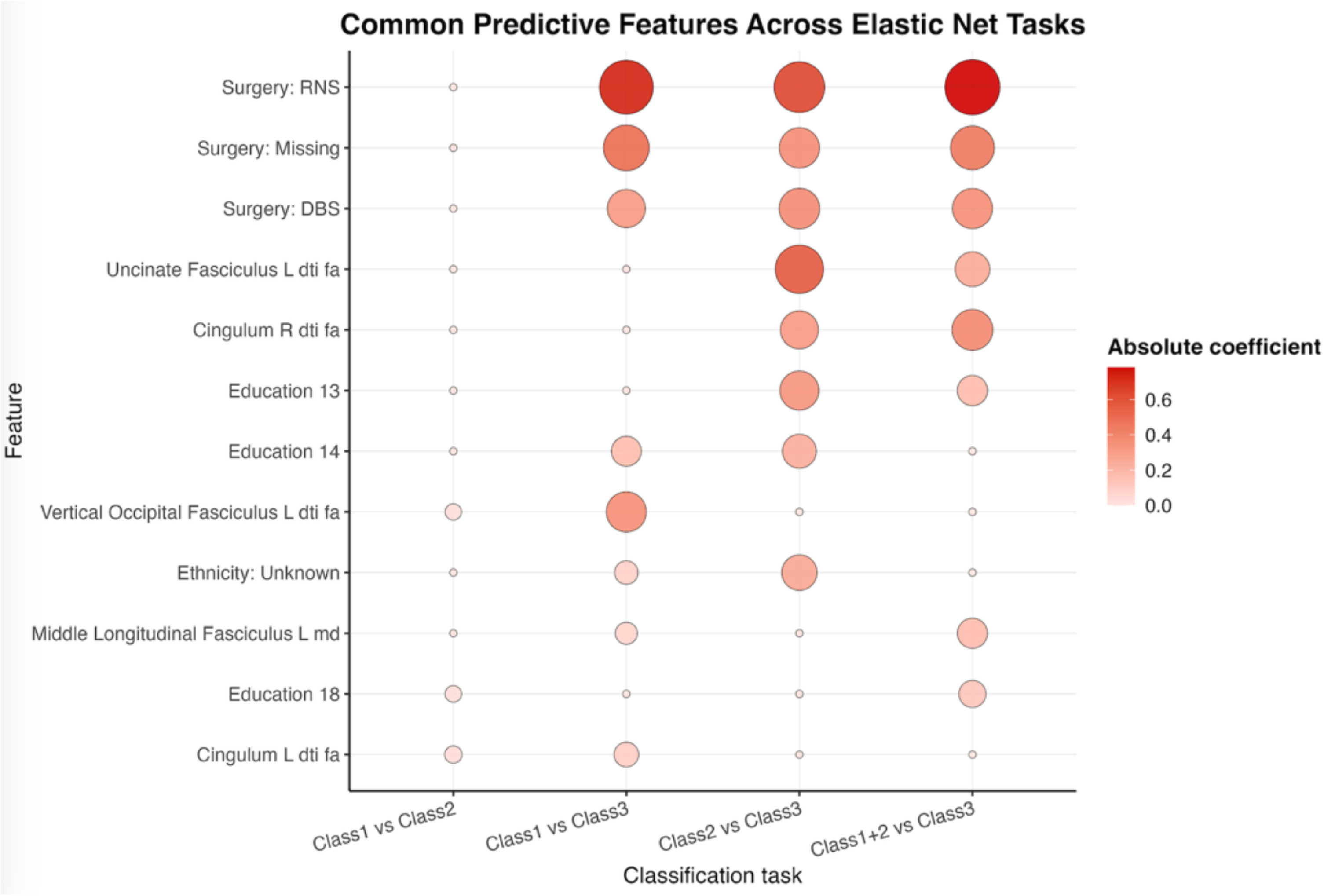
Common predictive features across elastic net classification tasks. Each row in the figure represents a feature, each column represents a classification task, and each circle corresponds to the absolute value of the elastic net regression coefficient for that feature within the respective model. Larger circles and darker red shading indicate stronger contribution to classification, whereas smaller and lighter circles indicate weaker effects.

## Discussion

Previous research examining the usefulness of DTI in epilepsy is relatively limited and has focused on using DTI to identify malformations of cortical development [27] and focal cortical dysplasia [28, 29]. To our knowledge, this is the first reported use of DTI-T to identify abnormal white matter features in epilepsy patients directly. Given that epilepsy is a network disease, these findings support the hypothesis that specific white matter fascicles would be impacted by seizure spread to distal regions of the brain [30].

Our aim was to determine if epilepsy source lateralization could be determined from a combination of DTI-T and clinical variables using a statistical machine learning model. Our hypothesis was largely supported, in that the model fits were good for discriminating unilateral from bilateral sources and right sided focal epilepsy from left, although with lower accuracy when comparing unilateral and bilateral cases. The models best separate unilateral (right or left) epilepsy from generalized/bilateral epilepsy. This could be particularly useful in situations where EEG suggests either a focal or a generalized pattern and leaves some ambiguity as to the ultimate diagnosis and the resulting treatment options. Clinicians commonly encounter this scenario, for example, in patients with early versive seizures. The sympatomatogenic zone for versive seizures resides in the frontal eye fields so frontal lobe epilepsy is a consideration with early versive seizures [31]. However, people with generalized epilepsy can also have early versive seizures [32]. EEG does not always differentiate between the two so DTI-T lateralization could remove the need for more invasive testing like sEEG when scalp EEG is not sufficient.

Our models indicated that right sided focal epilepsy was harder to discriminate from left sided focal epilepsy, with an AUC of 0.758. This is still a useful classification, but weaker than the model fits comparing with bilateral epilepsy. This weaker classification may be the result of the commonly observed pattern of seizures secondarily generalizing or propagating bilaterally, which could imply that white matter in both hemispheres would potentially be affected. However, this is beyond the scope of our initial study. More research on the direct impact of seizures on white matter integrity is needed.

In addition to DTI-T biomarkers providing a potential tool to aid in the localization of seizure foci or clarifying the diagnosis of generalized versus focal epilepsy, we hypothesize DTI-T has potential to improve understanding of how seizures spread between different cortical and subcortical regions. This could lead to considering therapeutic targets involving white matter as opposed to classic targeting of only gray matter. There is growing interest in white matter neuromodulation in epilepsy with recent studies demonstrating potential promise of this strategy in DRE patients. For instance, a small study examined responsive neurostimulation outcomes involving the parahippocampal white matter in patients with bilateral temporal lobe epilepsy, showing reduced seizure frequency by 40-50% in most patients [33]. While novel to epilepsy, targeting white matter structures has proved successful in other diseases, primarily in psychiatric conditions. For instance, individuals with treatment resistant depression have been successfully treated with deep brain stimulation (DBS) implantation targeting areas of white matter such as the superolateral medial forebrain bundle [34]. High frequency stimulation of the subcallosal cingulate white matter, an area containing the cingulum bundle and portions of the uncinate fasciculus, have likewise shown some efficacy in depression [35].

DTI, while a well-established neuroradiological technique for understanding structural organization of the white matter in the brain, is not a standard evaluation tool for epilepsy. MRI for pre-surgical planning is standard of care and is well-tolerated by patients. DTI images can be collected over a few minutes within the same MRI scanning session and requires no additional radiotracer injection or active engagement with functional cognitive tasks during scanning, making the addition of DTI imaging to standard MRI acquisition relatively straightforward. While previous DTI studies in epilepsy have focused on examining epileptic lesions [27–29], we hypothesize that there is clinically useful information for lateralizing epileptogenic source within the individual white matter tracts. We further propose that this information can be harnessed using a novel analytic technique, specifically, elastic net regularization analysis that can reveal patterns hidden within multivariate and correlated features. This additional structural information about an individual patient’s white matter, along with radiologic findings of lesions or cortical malformations on structural MRI, can enhance the functional approaches already in use for clinical evaluation, including EEG, MEG, PET, and fMRI.

Overall, our study indicates that DTI-T biomarker features analyzed with elastic net regularization can effectively distinguish epilepsy lateralization with a favorable balance between sensitivity and specificity, supporting its potential utility as an interpretable clinical decision-support tool.

## Limitations

One limitation of surgical epilepsy research is that there is no a priori way of identifying the exact EZ prior to therapeutic intervention. Although we believe it is reasonable to assume that the epileptogenic foci of those individuals who are seizure free after surgery were accurately classified, those who are not seizure free likely have more complicated seizure networks not targeted during surgery, or the surgical workup led to the wrong conclusion altogether. Our population used the epilepsy surgery conference consensus as the gold standard to classify the localization and lateralization of our sample. Given the constraints noted above, this is obviously problematic. A potential way to strengthen our model might be to include only individuals who were seizure free following surgery. However, that approach would automatically exclude those with more complex forms of epilepsy. Future research will need to focus on the predictive ability of our model to further elucidate its real-world efficacy.

Our ultimate goal is to incorporate DTI-T biomarker metrics into presurgical planning; however, we decided to include surgical decisions from our clinicians in the models presented in this paper, which would of course not be available on new cases. Across our paired classification tests, these surgical decisions were among the most important discriminating features when comparing unilateral and bilateral cases, alongside several DTI and clinical characteristics. As the EZ and surgical decisions in our data were determined without the DTI-T biomarkers included in our analysis, we believe this supports our hypothesis that DTI-T biomarkers carry clinically relevant information that is aligned with our current clinical understanding of epilepsy. More research on how the DTI-T biomarker metrics relate with other clinically relevant data and biomarkers should be sought.

It is also noteworthy that these data were generated from a single epilepsy center with a high volume of complex cases that aggressively evaluates patients for seizure freedom with surgical methods. The process of patient selection, characteristics, and outcome goals varies between centers, so the statistical models generated here would need to be validated before being used in clinical decision making.

Some potential next steps would be to examine how incorporating other pre-surgical evaluation data (i.e., EEG, PET, MEG) changes the model outcomes and using the models to predict seizure outcomes following surgery. One interesting new approach in machine learning models is to incorporate network structural information into classification models to improve predictive performance [36–38] . White matter tractography and region-to-region network relationships would be an excellent data source for this new machine learning network analysis approach.

## Conclusion

Our novel analytic approach, elastic net regularization of DTI data, suggests there is value in DTI-T biomarker metrics for distinguishing epilepsy lateralization, supported by the model alignment with previously made clinical decisions. The features highlighted by our models were largely in agreement with the clinical lateralization identified during our center’s surgical consensus conference, rather than the DTI data alone. However, these results should be interpreted cautiously as this is a preliminary biomarker validation and novel application that has yet to be thoroughly evaluated in the context of clinical decision making. Additional research is needed to understand whether information contained within normal appearing white matter can be used to improve surgical decision-making and outcomes for patients with epilepsy.

## Supporting information

Tabl S3

## List of Abbreviations

3T: 3 Tesla
ASM: Antiseizure Medications
AUC: Area Under the Receiver Operating Characteristic Curve
DBS: Deep Brain Stimulation
DNN: Deep Neural Network
DTI: Diffusion Tensor Imaging
DRE: Drug Resistant Epilepsy
EZ: Epileptogenic Zone
FA: Fractional Anisotropy
fMRI: Functional Magnetic Resonance Imaging
GBM: Gradient Boosting Machine
7T MRI: High-Field
SPECT: Ictal Single Photon Emission Computed Tomography
MRI: Magnetic Resonance Imaging
MPRAGE: Magnetization Prepared Rapid Acquisition Gradient Echo
MEG: Magnetoencephalography
MD: Mean Diffusivity
PET: Positron Emission Tomography
RF: Random Forest
ROC: Receiver Operating Characteristic
sEEG: Stereo Electroencephalogram
SVM: Support Vector Machine
VNS: Vagus Nerve Stimulator
vEEG: Video Electroencephalogram

## Acknowledgements

We humbly thank the patients who contributed their imaging and other data for this retrospective analysis. We also thank the Epilepsy team at the University of Kansas Health System who participated in the pre-surgical planning and consensus clinical conference diagnosis for these patients.

## Data availability statement

All data (group means and measures of dispersion) used in the present analyses are included in tables 1, 2, 3 and supplementary tables S1, S2.

## IRB statement

This study was approved by the University of Kansas Medical Center Human Research Protection Program, and all data were collected into the research database from retrospective chart review as part of an ongoing approved IRB Study (Approval # STUDY00160649) with waiver of consent in accordance with the Declaration of Helsinki. All patients at the University of Kansas Medical Center sign a general consent for retrospective chart review.

## Ethical publication statement

We confirm that we have read the Journal’s position on issues involved in ethical publication and affirm that this report is consistent with those guidelines.

## Permission to reproduce material from other sources

N/A

## Clinical trial registration

N/A

## Author contributions

Conceptualization: RJL, JH, YL, CMP; Data Curation: MP, XC, CMP; Formal Analysis: XC, JH, YL, MA; Funding Acquisition: RJL, CMP; Investigation: MP, CMP; Methodology: XC, JH, YL, MA; Project Administration: RJL, CMP; Resources: MF, PL, CMP; Software: XC, JH, YL, MA; Supervision: JH, YL; Validation: XC, JH, YL, MA; Visualization: XC, MA; Writing – original draft: RJL, MP, XC, CMP; Writing – review & editing: JH, YL, MA, MF, PL

## Generative AI statement

No generative AI (LLM) was used in the writing of this manuscript.

## Supplemental information

Table S1. GREENBEAN Checklist.

Table S2. Fractional anisotropy (FA) from white matter tracts included in the analysis.

Table S3. Mean diffusivity (MD) from white matter tracts included in the analysis.

Figure S1. Visualization of the variability within the whole brain A. fractional anisotropy (FA) and B. mean diffusivity (MD) across participants.

