## Supplementary material for "Epilepsy Lateralization by Elastic Net Regularization of a Diffusion Tensor Imaging Biomarker using Machine Learning": Table S2

Table S2. Fractional anisotropy (FA) from white matter tracts included in the analysis.

| Tract | Mean | SD | Missing |
|---|---|---|---|
| **FA** | | | |
| Whole Brain | 0.41 | 0.02 | 0 |
| Arcuate Fasciculus Left | 0.44 | 0.03 | 4 |
| Arcuate Fasciculus Right | 0.43 | 0.04 | 45 |
| Cingulum Left | 0.37 | 0.04 | 0 |
| Cingulum Right | 0.38 | 0.04 | 0 |
| Extreme Capsule Left | 0.38 | 0.03 | 80 |
| Extreme Capsule Right | 0.40 | 0.03 | 58 |
| Frontal Aslant Left | 0.39 | 0.03 | 3 |
| Frontal Aslant Right | 0.39 | 0.03 | 1 |
| Inferior Fronto Occipital Fasciculus Left | 0.45 | 0.03 | 6 |
| Inferior Fronto Occipital Fasciculus Right | 0.46 | 0.03 | 5 |
| Inferior Longitudinal Fasciculus Left | 0.42 | 0.04 | 0 |
| Inferior Longitudinal Fasciculus Right | 0.43 | 0.03 | 0 |
| Middle Longitudinal Fasciculus Left | 0.39 | 0.04 | 81 |
| Middle Longitudinal Fasciculus Right | 0.40 | 0.03 | 21 |
| Parietal Aslant Left | 0.38 | 0.03 | 0 |
| Parietal Aslant Right | 0.39 | 0.03 | 0 |
| Superior Longitudinal Fasciculus (1) Left | 0.38 | 0.03 | 0 |
| Superior Longitudinal Fasciculus (1) Right | 0.41 | 0.04 | 0 |
| Superior Longitudinal Fasciculus (2) Left | 0.39 | 0.03 | 0 |
| Superior Longitudinal Fasciculus (2) Right | 0.41 | 0.04 | 2 |
| Superior Longitudinal Fasciculus (3) Left | 0.38 | 0.04 | 0 |
| Superior Longitudinal Fasciculus (3) Right | 0.41 | 0.04 | 0 |
| Uncinate Fasciculus Left | 0.33 | 0.03 | 0 |
| Uncinate Fasciculus Right | 0.35 | 0.03 | 0 |
| Vertical Occipital Fasciculus Left | 0.37 | 0.03 | 0 |
| Vertical Occipital Fasciculus Right | 0.38 | 0.03 | 0 |
| Cortico-Striatal Pathway Left | 0.40 | 0.03 | 0 |
| Cortico-Striatal Pathway Right | 0.42 | 0.03 | 0 |
| Corticothalamic Pathway Left | 0.41 | 0.02 | 0 |
| Corticothalamic Pathway Right | 0.42 | 0.03 | 0 |
| Fornix Left | 0.26 | 0.04 | 13 |
| Fornix Right | 0.27 | 0.03 | 34 |
| Cerebellum Left | 0.19 | 0.02 | 0 |
| Cerebellum Right | 0.19 | 0.02 | 0 |

Abbreviations: SD – standard deviation, FA – fractional anisotropy
