## Supplementary material for "Epilepsy Lateralization by Elastic Net Regularization of a Diffusion Tensor Imaging Biomarker using Machine Learning": Tabl S3

Table S3. Mean diffusivity (MD) from white matter tracts included in the analysis.

| Tract | Mean | SD | Missing |
|---|---|---|---|
| **MD** | | | |
| Whole Brain | 0.88 | 0.04 | 0 |
| Arcuate Fasciculus Left | 0.78 | 0.06 | 4 |
| Arcuate Fasciculus Right | 0.76 | 0.05 | 45 |
| Cingulum Left | 0.83 | 0.08 | 0 |
| Cingulum Right | 0.81 | 0.08 | 0 |
| Extreme Capsule Left | 0.83 | 0.07 | 80 |
| Extreme Capsule Right | 0.82 | 0.08 | 58 |
| Frontal Aslant Left | 0.77 | 0.06 | 3 |
| Frontal Aslant Right | 0.75 | 0.06 | 1 |
| Inferior Fronto Occipital Fasciculus Left | 0.85 | 0.06 | 6 |
| Inferior Fronto Occipital Fasciculus Right | 0.83 | 0.05 | 5 |
| Inferior Longitudinal Fasciculus Left | 0.87 | 0.09 | 0 |
| Inferior Longitudinal Fasciculus Right | 0.84 | 0.08 | 0 |
| Middle Longitudinal Fasciculus Left | 0.82 | 0.08 | 81 |
| Middle Longitudingal Fasciculus Right | 0.80 | 0.06 | 21 |
| Parietal Aslant Left | 0.80 | 0.07 | 0 |
| Parietal Aslant Right | 0.80 | 0.08 | 0 |
| Superior Longitudinal Fasciculus (1) Left | 0.78 | 0.07 | 0 |
| Superior Longitudinal Fasciculus (1) Right | 0.76 | 0.08 | 0 |
| Superior Longitudinal Fasciculus (2) Left | 0.77 | 0.07 | 0 |
| Superior Longitudinal Fasciculus (2) Right | 0.76 | 0.08 | 2 |
| Superior Longitudinal Fasciculus (3) Left | 0.79 | 0.08 | 0 |
| Superior Longitudinal Fasciculus (3) Right | 0.77 | 0.09 | 0 |
| Uncinate Fasciculus Left | 0.89 | 0.07 | 0 |
| Uncinate Fasciculus Right | 0.84 | 0.08 | 0 |
| Vertical Occipital Fasciculus Left | 0.78 | 0.06 | 0 |
| Vertical Occipital Fasciculus Right | 0.79 | 0.04 | 0 |
| Cortico-Striatal Pathway Left | 0.80 | 0.05 | 0 |
| Cortico-Striatal Pathway Right | 0.80 | 0.05 | 0 |
| Corticothalamic Pathway Left | 0.79 | 0.07 | 0 |
| Corticothalamic Pathway Right | 0.78 | 0.06 | 0 |
| Fornix Left | 1.58 | 0.23 | 13 |
| Fornix Right | 1.53 | 0.22 | 34 |
| Cerebellum Left | 0.91 | 0.09 | 0 |
| Cerebellum Right | 0.92 | 0.10 | 0 |

Abbreviations: SD – standard deviation, MD – mean diffusivity
