## Supplementary figures and images for "Epilepsy Lateralization by Elastic Net Regularization of a Diffusion Tensor Imaging Biomarker using Machine Learning"

### Figure S1

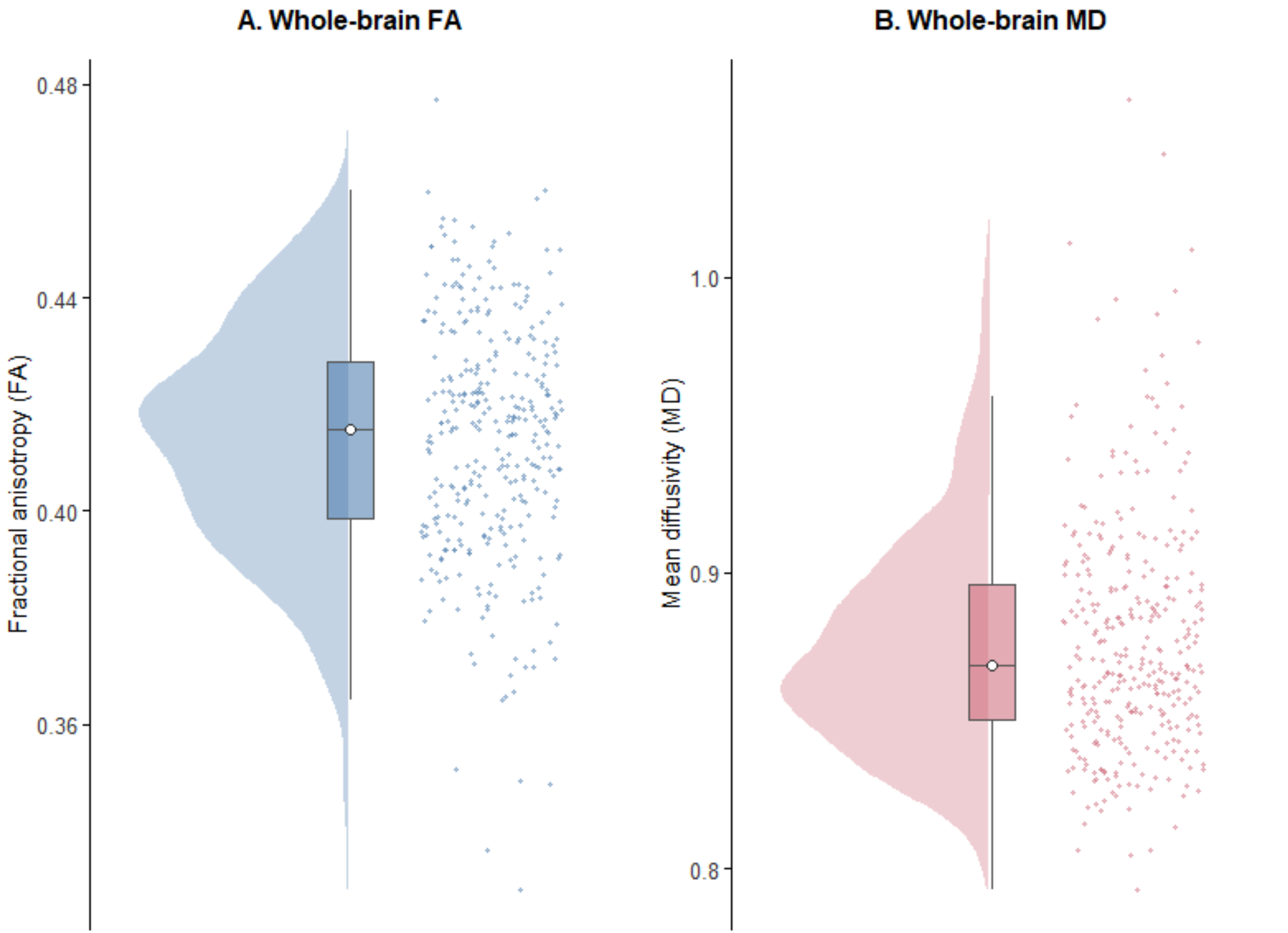
A. Whole-brain FA
B. Whole-brain MD
Fractional anisotropy (FA)
0.48
0.44
0.40
0.36
Mean diffusivity (MD)
1.0
0.9
0.8
